# QUALITY CONTROL OF *STAPHYLOCOCCUS AUREUS* IN RIO GRANDE AND MENONITA CRIOLLO CHEESES FROM THE ALEJO CALATAYUD MARKET

**DOI:** 10.1101/2022.03.01.22271584

**Authors:** Choque Rojas Litze Abigayl, Fuentes Condori Rosy, López Vargas Janethe Vania, Pardo Aranibar Carlos, Rojas Nogales Magali Dian

## Abstract

**Introduction:** The *Staphylococcus aureus* count is one of the indicators of the microbiological quality of cheeses according to the parameters of the Bolivian Standard 32004, since this bacterium can cause food poisoning.

**Objective:** **To** determine the presence and quantity of *Staphylococcus aureus* in Rio Grande and Menonita Creole cheeses in the Alejo Calatayud Market in the city of Cochabamba during the month of July 2019 according to parameters established by Bolivian Standard 32004.

**Methods:** Eight samples taken from different cheeses randomly selected from the market were analyzed and subjected to quantitative analysis of CFU/g of *Staphylococcus aureus* in three different dilutions for each sample, giving a total of 24, 13 were discarded due to impossibility of counting and the remaining samples were subjected to confirmatory tests. The parameters established in the NB32004 were used for the qualification of the results of analysis and tests.

**Results:** In 100% of the samples (8 of 8), the presence of S. *aureus* bacteria was found in quantities higher than the acceptable CFU/g, determining as deficient the hygienic-sanitary quality of the ripened Creole cheeses sold in the Alejo Calatayud Market.

**Conclusion:** Creole cheeses marketed in the Alejo Calatayud Market have a high degree of contamination by *S. aureus* and therefore do not comply with the parameters of NB32004 and the population is exposed to food poisoning.

## INTRODUCTION

*Staphylococcus aureus inhabits* the upper respiratory tract, nose and throat, as well as the skin surface of 30% of the healthy human population and causes various diseases in humans and animals (1).

In the case of cheese contamination can occur in the different links of the production chain, the deficiencies in its handling by those responsible for its preparation, determine problems in public health, mainly in developing countries (2).

In Spain, studies on *S. aureus* reflect a prevalence of infection from 1.5 % of infected persons in 2010, up to 18 - 23% in 2013, in certain periods raising these figures up to 40%. In Venezuela, a study was carried out to quantitatively determine the presence of *S. aureus* in white cheese sold in the city of Mérida-Venezuela. A total of 72 samples were analyzed and S. aureus was detected in 69.44% of the cheeses; loads above 10 3CFU/g in 41.67% of the samples and above 105 in 8.34%. When comparing the results with the microbiological requirements demanded for this type of product, it is concluded that there is a great hygienic deficiency and represents a latent danger as a vehicle of staphylococcal intoxication for the consumer (4).

According to Yugcha Silvana Pamela: “*S. aureus* infections have been reported in pasteurized cheeses in different parts of Latin America. In Mérida (Venezuela) 64.44% were obtained; in Lima (Peru) 53.4%; in Corrientes (Argentina) 45%; in Tabasco (Mexico) 11.8% and in Ecuador in a study conducted in November and December 2015, the prevalence of *S. aureus* in cheeses was determined to be 83.33%.”(5)

Likewise, a study carried out in Santa Cruz de la Sierra, October 11 to 13, 2018 by the Faculty of Veterinary Sciences, Universidad Autónoma Gabriel René Moreno determined the hygienic and sanitary quality of cheeses from the Valle Grande markets, demonstrating a high degree of contamination. Of the outbreaks of intoxication that occur, on average, 20% are due to the consumption of food contaminated with bacteria of the *Staphylococcus* genus and, mainly, by the aureus species.

Therefore, the present study seeks to determine if the count of positive *Staphylococcus* coagulase in these cheeses is within the parameters established in the Bolivian Standard NB 32004, and thus consider whether it is suitable for human consumption, since this food is consumed daily by the entire population directly or as an ingredient in different types of meals, highlighting that the Alejo Calatayud Market in the city of Cochabamba is a very busy supply center.

## MATERIALS AND METHODS

The present study corresponds to an observational, analytical, cross-sectional research. In Mennonite and Rio Grande cheeses of the Alejo Calatayud Market, selected by means of random convenience sampling, based on inclusion and exclusion criteria.

Cheeses sold in fixed stalls in the market were selected for inclusion, excluding cheeses sold in other places.

The instruments used in the collection of samples were stainless steel knife and plastic bags or sacks with hermetic seal. The culture media used were Petri dishes with Baird Parker agar and others with Mannitol salt agar. The solutions, reagents and indicators were Gram stain reagents (crystal violet, lugol, acetone alcohol, safranin, water) and hydrogen peroxide.

The materials and equipment used were sterile graduated pipettes of 1 mL with cotton plug (one for each dilution), sterile Pasteur pipettes, sterile graduated pipettes of 10 mL with cotton plug, sterile glass rods of 3.5 mm diameter approx, sterile sterile utensils: knives, spoons, tweezers, bacteriological handle, microscope slides, optical microscope, balance, oven for sterilizing glassware, autoclave, boiling water bath and incubator.

### Laboratory Procedure

#### Sample transport

The samples were placed in first-use polyethylene bags and then were Processed immediately within 2 hours after sampling.

#### Preparation and dilution of sample homogenates (fig. 3)

First we cut a small amount of each sample, we crushed it very well in a mortar, we weighed only 10 g in the balance and introduced it into a previously sterilized flask for each type of cheese, then we added 90 ml of physiological solution, we had to shake the samples manually because we did not have the necessary equipment. Then we made the decimal dilutions in 1:10, 1:100 and 1: 1000 in sterilized test tubes obtaining 24, we placed 0.4ml of each dilution for each Petri dish with Baird Parker agar, extending the inoculated volume with a sterile glass rod, in the form of “L”. we waited between 5 and 10 min. approx. for the inoculum to be absorbed and then we took to incubate during 24-48 h. After the incubation time, the characteristic colonies of this microorganism were observed as black, circular, shiny, smooth colonies, showing an opaque circular zone and a clear halo around it due to the presence of tellurite, lithium chloride and glycine. Their presence is indicated by a dark appearance due to the reduction of tellurium.

**Fig. 1.**
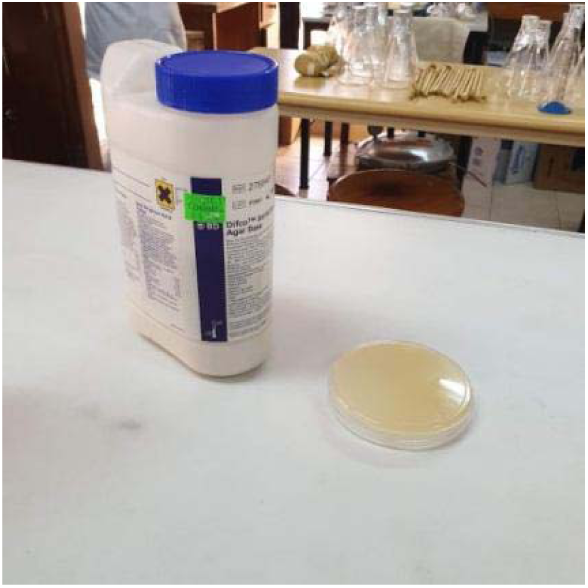
Baird Parker Agar.

**Fig. 2.**
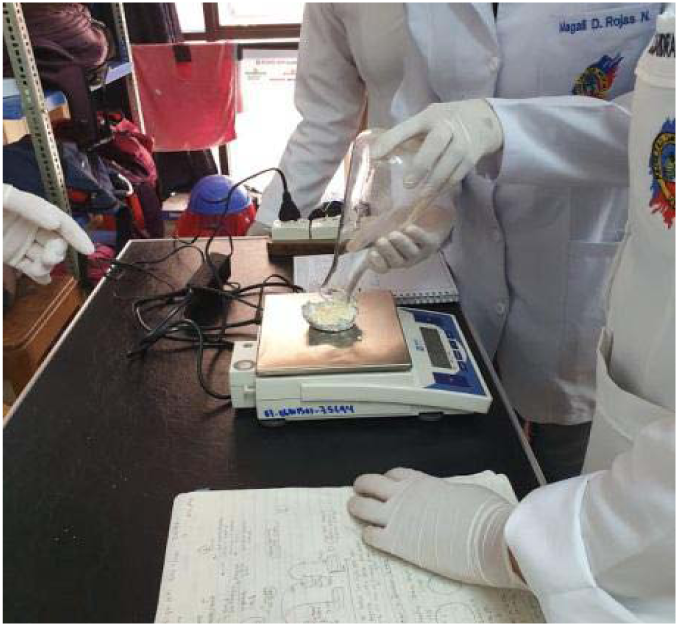
Weighing: 10 grams of each cheese.

**Fig.3.**
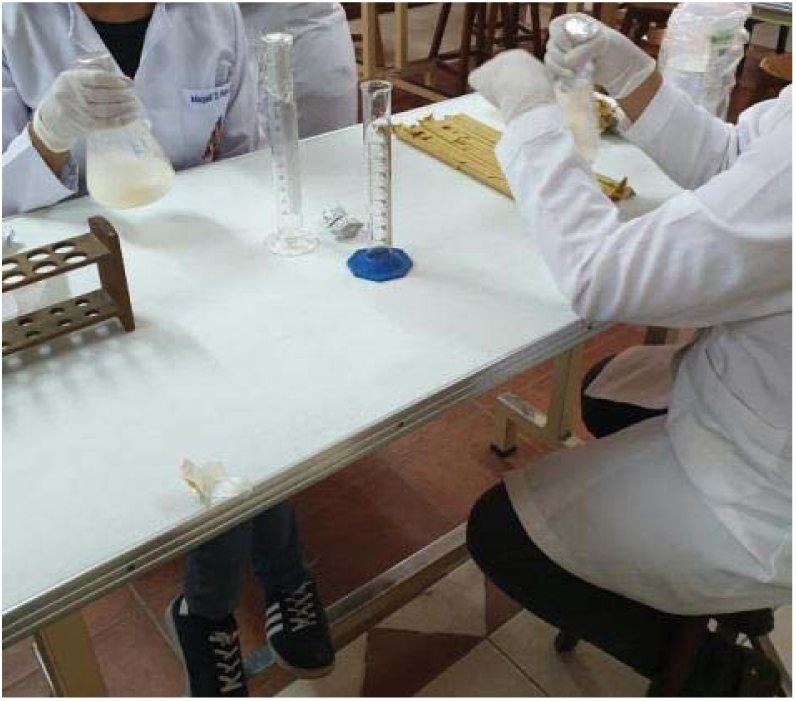
Cheese homogenization with sterile water.

#### Culture of Samples in Mannitol Salt Selective Agar

In the laboratory, the selected colonies of the Baird Parker agar were sterile seeded on mannitol-salt selective agar. Each of the samples was streaked with the help of a bacteriological loop, covering the entire surface of the agar, at three different angles. The plates were then incubated at 37°C for 48 hours. The plates were read 24 and 48 hours after seeding. The selected CFU were yellowish-white, even orange-colored colonies. (Fig. 6)

**Fig. 4.**
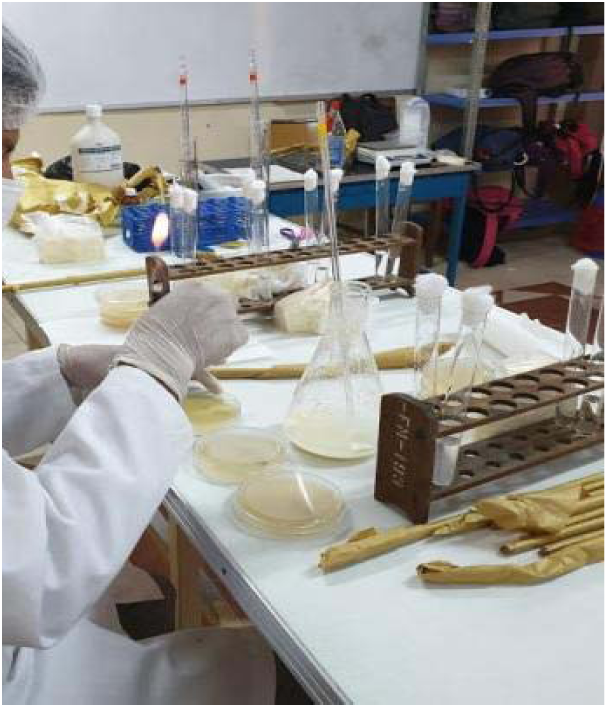
Seeding on Baird Parker Agar.

**Fig. 5.**
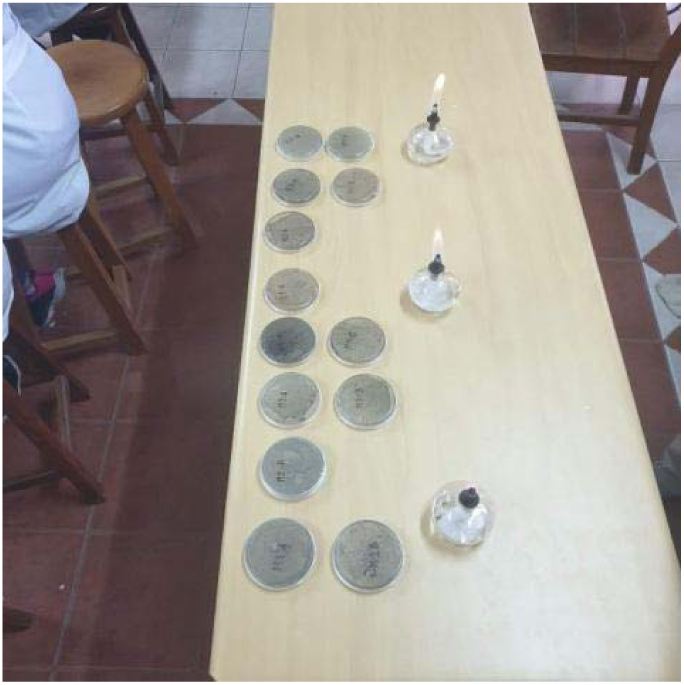
Incubation for 24 hours.

**Fig. 6.**
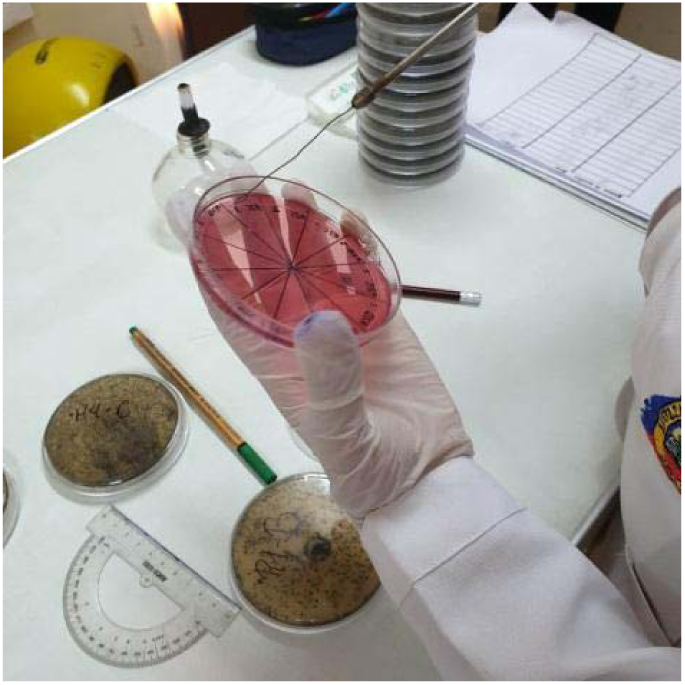
Mannitol salt test.

#### Biochemical tests

##### * Catalase test

With a bacteriological loop, we took an inoculum of Baird Parker agar after its incubation, and placed it on a slide, adding one or two drops of 30% hydrogen peroxide, mixed perfectly well and observed the results. The formation of bubbles, the test was considered positive. (fig. 9)

**Fig.7.**
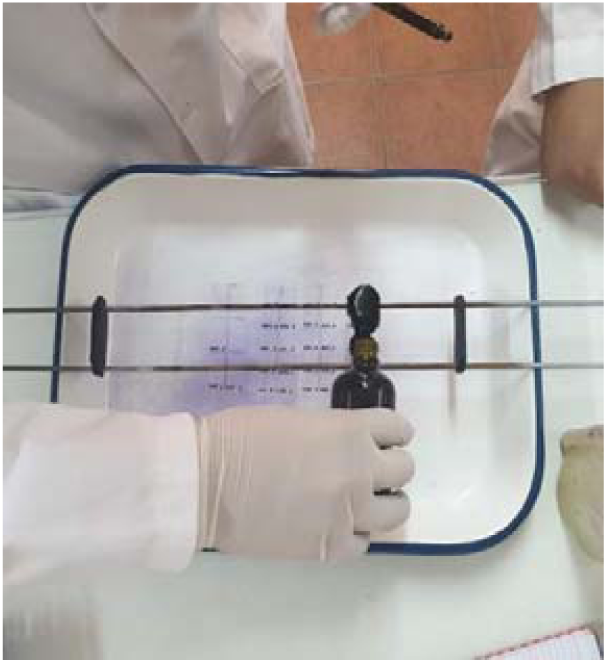
Gram staining procedure.

**Fig. 8.**
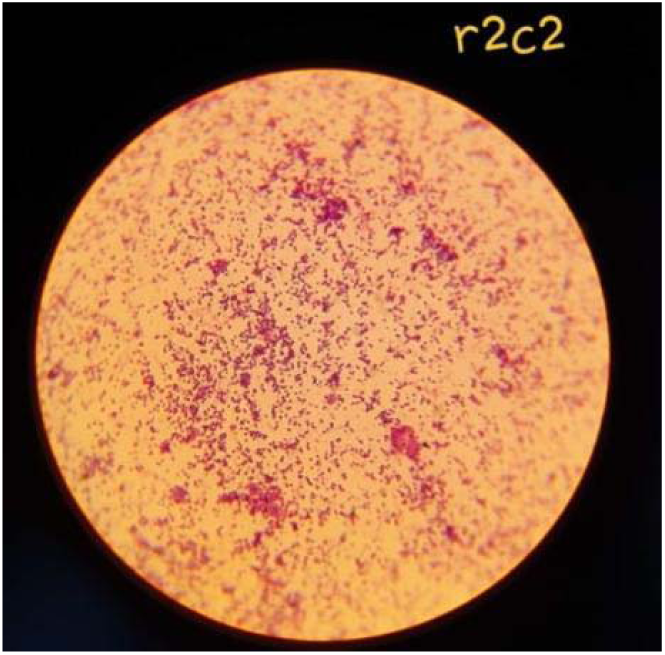
Plates viewed under the microscope.

**Fig. 9.**
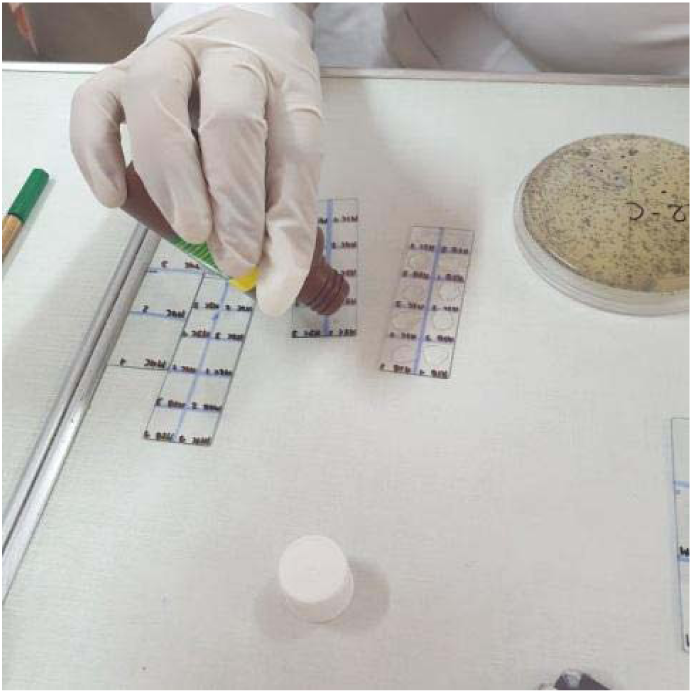
Catalase test.

**Fig. 10.**
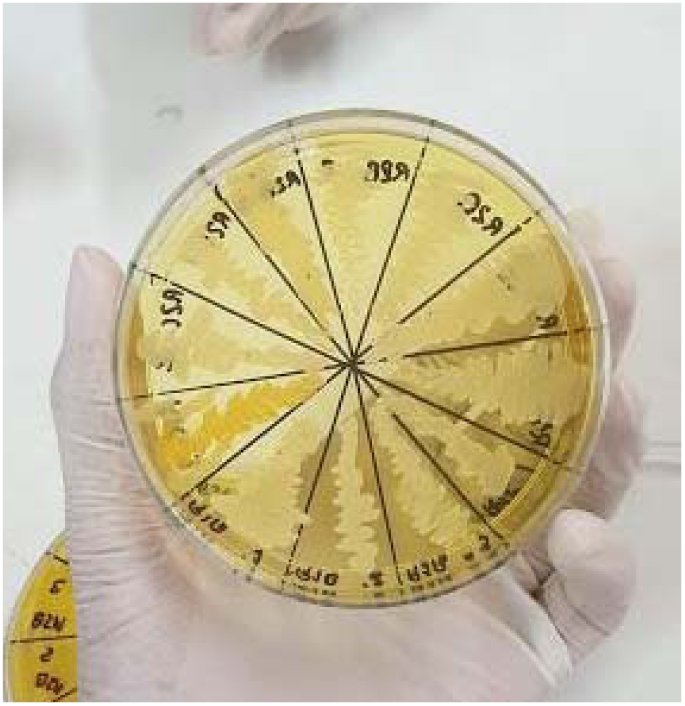
Positive mannitol test.

##### Gram stain

After the catalase test, Gram staining was performed. We covered the slides with crystal violet, leaving it to act for 1min, washed with running water, added lugol and left it to act for 1min, washed again, decolorized with acetone alcohol for 30 minutes, washed and added safranin for 1 to 3 min, washed, left to dry and observed under the microscope.

## RESULTS

The presence of *Staphylococcus aureus* was demonstrated in 22 samples out of 24 (13 samples were uncounted and all of them were considered positive, and the other 11 were worked) as well as quantifying the CFU/gr of each sample and comparing with the parameters established by the Bolivian standard 32004, it was concluded that none of them is within the established range, all of them exceed the range. (Table 4)

**TABLE 1.**
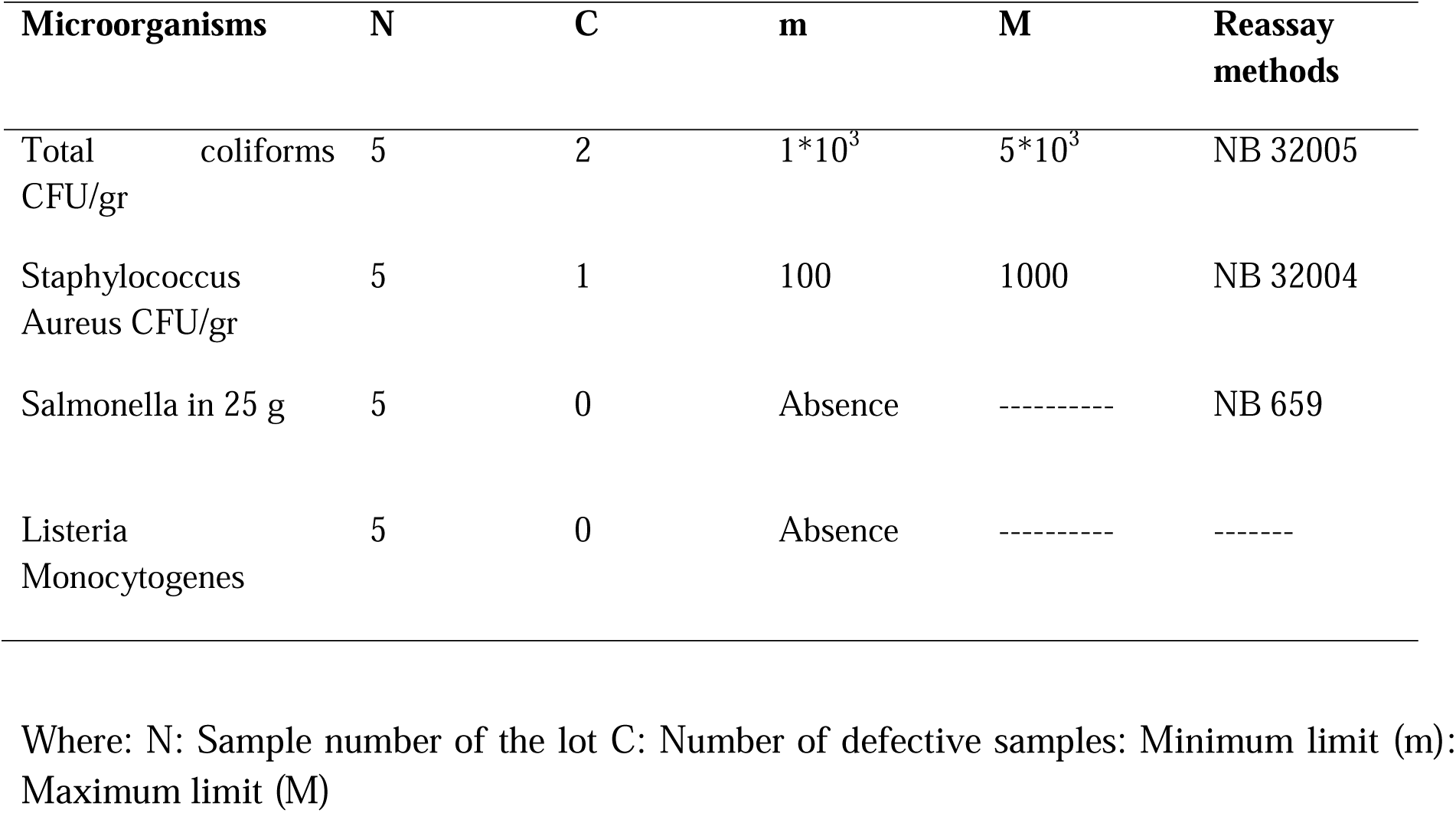
BOLIVIAN STANDARD NB32004.

**TABLE2.**
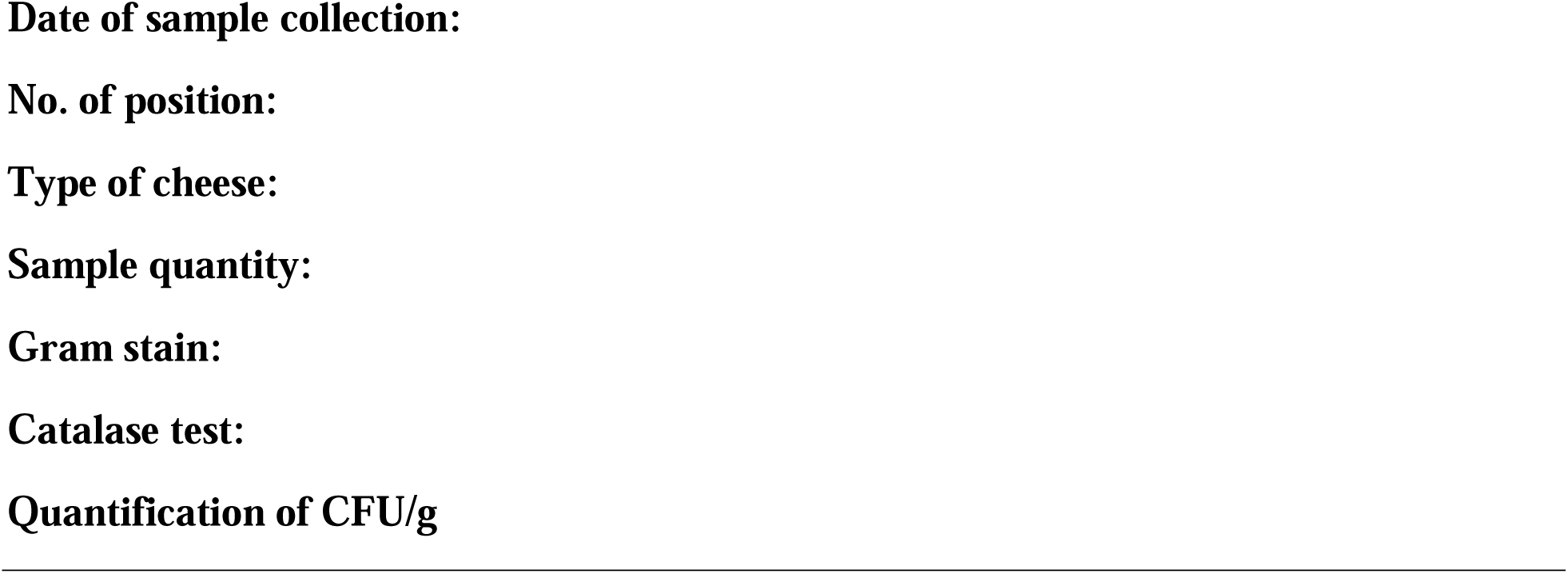
DATA COLLECTION FORM.

**TABLE. 3.**
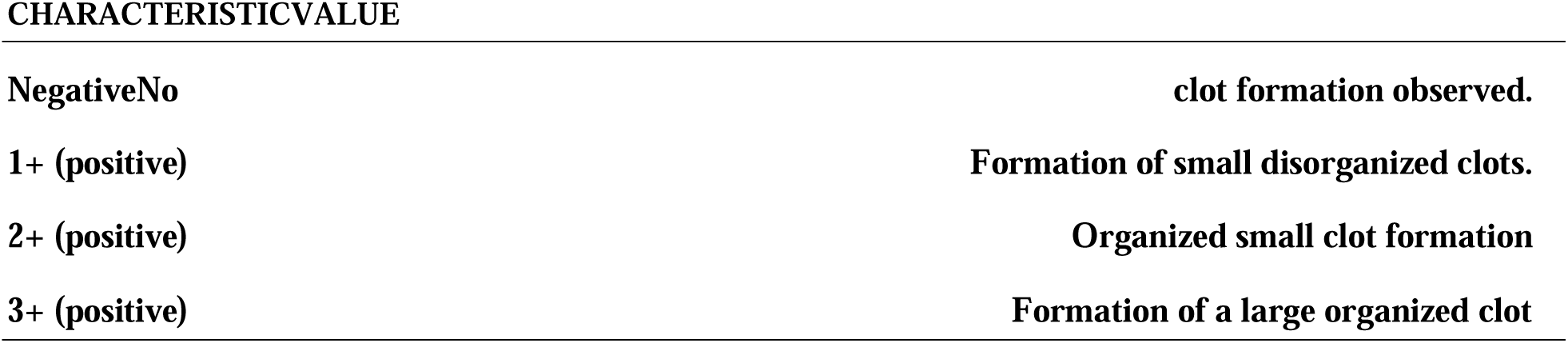
Interpretation of reactions in the coagulase test.

**TABLE 4.**
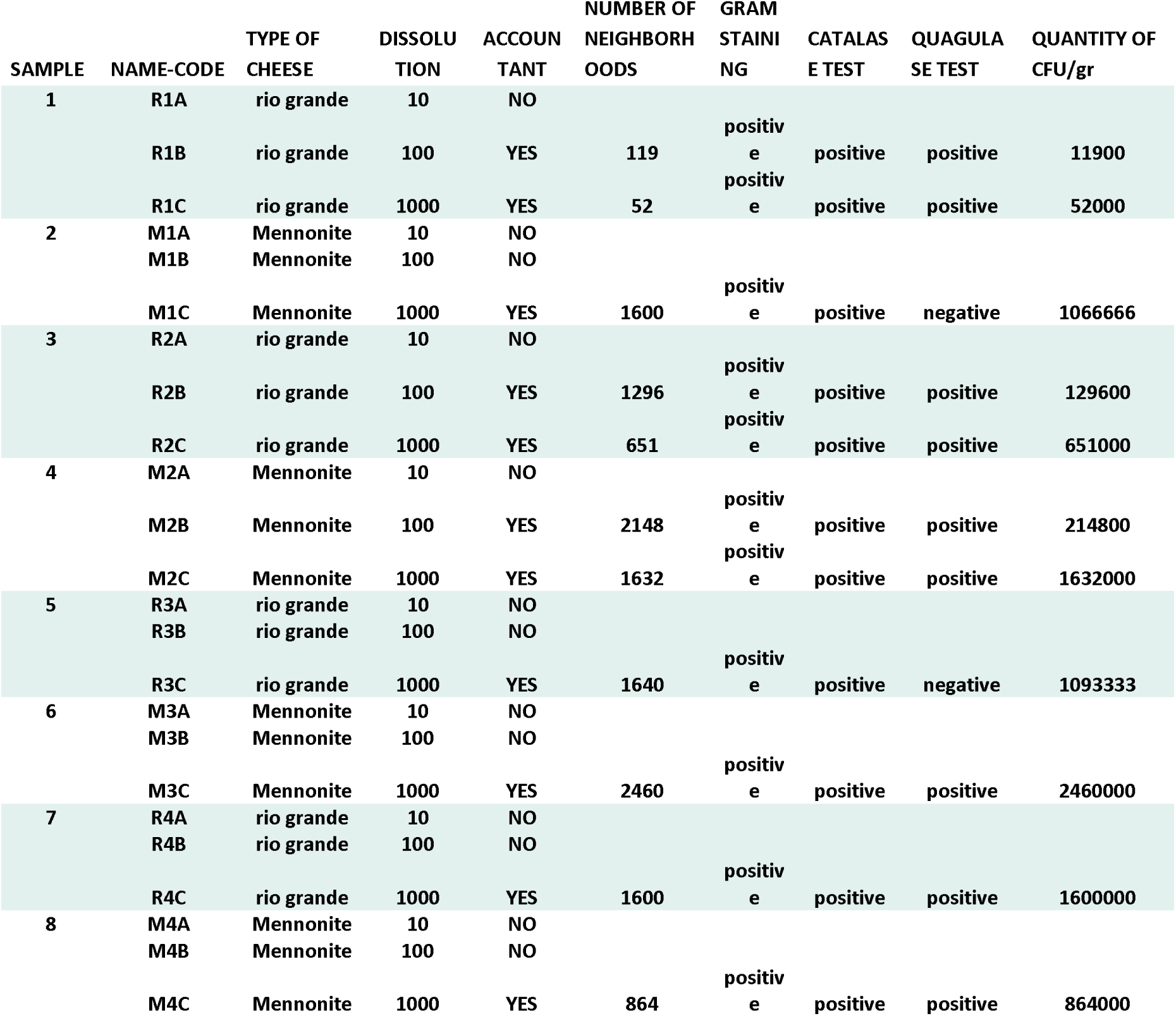
DATA TABULATION.

## DISCUSSION

Our work shows a higher presence of *S. aureus bacteria* in Rio Grande and Mennonite Creole cheeses from Alejo Calatayud Market in the town of Cochabamba during the month of July 2019, in comparison with a work carried out in Santa Cruz de la Sierra, October 11-13, 2018 by the Faculty of Veterinary Sciences, Gabriel René Moreno Autonomous University with the purpose of determining the hygienic and sanitary quality of cheeses from Valle Grande markets(11), this fact could represent an important risk to the health of consumers seriously affecting marketing and their interest in artisanal cheeses. This totally contrasts with the results of a study in the State of Tabasco by Borbolla-Sala, and with another study in Chiuata de Tapia in which they found levels of *S. aureus* within the limits allowed by Mexican Standards(12).

The presence of *S. aureus* in artisanal cheeses is related to the initial concentration in the raw milk, in the steps of the manufacturing process or finally in the handling of the product by those who market it. On the other hand, it has been indicated that a high number of microorganisms, although indicative of poor handling, is not sufficient to link the product with the risk of food poisoning, and its capacity to produce whole toxins must be demonstrated (13).For obvious reasons, the maximum microbiological limits allowed should be below the values indicated above in order to ensure a safe product, thus avoiding samples with *S. aureus*, between 100 000 and above 1 000 CFU/g represent an imminent danger for consumers, as they have a greater possibility of containing enterotoxins, since it has been corroborated that foods involved in intoxications contain high populations of the microorganism (14).a possibility of its multiplication during the distribution and commercialization of cheese.

For our analysis we used the bacteriological method of plate counting, with subsequent confirmation of suspicious colonies, which were confirmed by their submission to the coagulase and catalase tests “POSITIVE” compared with the parameters of the NB 32004 standard. It should be noted that this method is easy to perform, the reagents used are easy to prepare, accessible and available for work in any laboratory for food microbiology, although its main disadvantage is the time required for reading the results.

## CONCLUSIONS

The Creole cheeses sold in the Alejo Calatayud Market have a high degree of contamination by *S. aureus and therefore do* not comply with the parameters of NB32004 and the population is exposed to food poisoning.

## RECOMMENDATIONS

For cottage industry producers to have greater control over the quality of the cheese marketed as well as over the personnel involved in the production of the product. Verify that the raw milk for the production of creole cheeses arrives at the cottage industry in optimal transport conditions, in clean containers, refrigerated and from healthy animals.

## COMMUNITY OUTREACH

It is important to carry out awareness-raising activities among the vendors that sell this product, reaching an agreement to implement prevention and control strategies, which will be achieved with the support of multidisciplinary groups and, fundamentally, by educating the population about food safety.

Research on the degree of contamination of the cheeses is recommended for future work.

## ANNEXES

**Table 1.** Presence *of staphylococcus aureus in the samples*.

|  |  | Mennonite | rio grande | total |
| --- | --- | --- | --- | --- |
| S.AUREUS | POSITIVE | 11 | 11 | 22 |
|  | ANOTHER STAPHYLO | 1 | 1 | 2 |
| Total |  | 12 | 12 | 24 |

Source: own elaboration

**Table 2.**
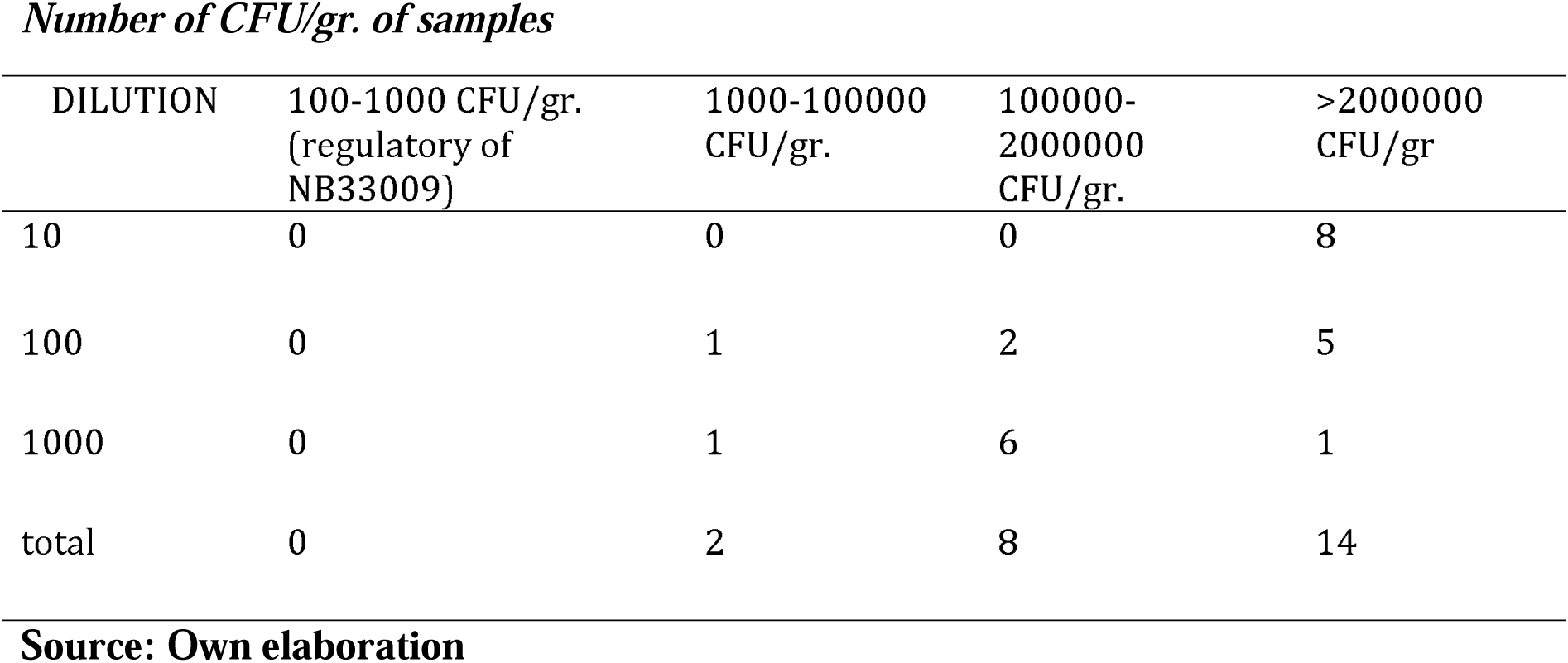
Number of CFU/gr. of samples.

## Data Availability

All data produced in the present study are available upon reasonable request to the authors

